# Potential policy interventions for slowing the spread of artemisinin-resistant *pfkelch* R561H mutations in Rwanda

**DOI:** 10.1101/2022.12.12.22283369

**Authors:** Robert J. Zupko, Tran Dang Nguyen, J. Claude S. Ngabonziza, Michee Kabera, Haojun Li, Thu Nguyen-Anh Tran, Kien Trung Tran, Aline Uwimana, Maciej F. Boni

**Affiliations:** Center for Infectious Disease Dynamics, Department of Biology, Pennsylvania State University, University Park, PA, USA; Malaria and Other Parasitic Diseases Division, Rwanda Biomedical Centre (RBC), Kigali, Rwanda; Research, Innovation and Data Science Division, Rwanda Biomedical Center (RBC), Kigali, Rwanda; Department of Clinical Biology, University of Rwanda, Kigali, Rwanda; Department of Computer Science, Columbia University, New York, NY, USA; Centre for Tropical Medicine and Global Health, Nuffield Department of Medicine, University of Oxford, Oxford, UK

**Keywords:** malaria, drug resistance, kelch13, artemisinin

## Abstract

Artemisinin combination therapies (ACTs) are highly effective at treating uncomplicated *Plasmodium falciparum* malaria. However, the emergence of a novel *pfkelch13* R561H mutation in Rwanda, with associated delayed parasite clearance, suggests that drug policy interventions are needed to delay the fixation and slow the spread of this mutation. Using a spatial, stochastic, individual-based model calibrated and validated for the Rwanda’s malaria epidemiology, we evaluate seventeen strategies aimed at minimizing treatment failures and delaying the spread of R561H. The primary measures evaluated are projected treatment failures and R561H allele frequency over three, five, and ten years. Lengthening courses of treatment, deploying multiple first-line therapies, and custom rotation strategies all provide a benefit when compared to the status quo. The best intervention options, five years into the future, result in slower spread of R561H (0.16 allele frequency difference) and absolute treatment failure counts that are 44% lower than projected under the status quo.

## Introduction

The introduction of artemisinin combination therapies (ACTs) has been instrumental in reducing the burden of *Plasmodium falciparum* malaria; however, the continued evolution of drug resistance by malaria parasites has the potential to undermine these advancements. Since the first appearance of artemisinin resistance in Cambodia in the 2000s [1,2], the spread of molecular markers associated with artemisinin resistance has largely been concentrated in Southeast Asia [3,4]. However, the *de novo* appearance of confirmed markers for artemisinin resistance in Rwanda [5] and Uganda [6,7] signals the need for interventions to be considered in the African context.

The ACT artemether-lumefantrine (AL) was adopted by Rwanda as the first-line therapy for uncomplicated falciparum malaria in 2006 as part of a comprehensive national strategic plan for malaria control [8]. Since adoption of AL, the *pfkelch13* R561H mutation has emerged and been validated as a marker for partial artemisinin resistance in samples collected as part of clinical drug efficacy studies between 2012 - 2015 [3]. 561H is associated with a delayed clearance half-life of 7.2 hours (IQR: 6.1 – 7.7 hours), similar to that of the 580Y mutation that emerged in Cambodia, and is associated with delayed clearance of the parasite [7,9]. Following the original identification of 561H in Gasabo, Rwanda [6], recent studies have found 561H in additional districts, with recent allele frequency measurements ranging from 0.045 to 0.219 [7,10–12]. These findings indicate that drug policy interventions are now needed to delay the fixation of 561H within the local *P. falciparum* population, and to reduce the impact of treatment failures due to artemisinin resistant parasites.

As of November 2022, WHO recommends six ACTs for the treatment of uncomplicated *P. falciparum* malaria: AL, artesunate-amodiaquine (ASAQ), artesunate-mefloquine (ASMQ), dihydroartemisinin-piperaquine (DHA-PPQ), artesunate/sulfadoxine–pyrimethamine (AS+SP), and artesunate-pyronaridine (AS-Pyr) [13]. Within sub-Saharan Africa (SSA) the predominant therapies deployed are AL and ASAQ, with AS+SP sometimes used for seasonal malaria chemoprophylaxis [14]. While ASMQ and DHA-PPQ are recommended by WHO, they see limited use in SSA with AL and ASAQ being preferred [15]. As such, SSA faces a constrained drug landscape that requires national drug policy interventions be balanced between delaying drug resistance evolution by the parasite – leading to increased drug failures over the long term – and ensuring that therapies currently given are highly efficacious.

Studies detecting the mutant 561H allele indicate both increasing allele frequency and geographic spread; however, there may still be a window of opportunity to delay the fixation of 561H in Rwanda and avert high numbers of treatment failures. Accordingly, we examined a range of possible drug policy interventions, the majority of which utilize existing therapies, and their ability to slow down 561H evolution and reduce long-term treatment failures. These include the replacement of the existing first-line therapy, introduction of multiple first-line therapies (MFT), and lengthening the dosing schedule for AL from a 3-day course of treatment to up to five days of treatment with AL in accordance with previous clinical trials [16–18]. Additionally, a more logistically complicated strategy of drug rotation, along with an early exploration of the possible impact of triple artemisinin combination therapies (TACTs) are included.

## Results

To provide a common point of comparison for policy interventions a baseline (or status quo) scenario was run (*n* =100 replicates) in which no interventions are implemented. Our spatially calibrated model – in good agreement with the 2017 Malaria Atlas Project projections for Rwanda’s malaria prevalence (i.e., the *P. falciparum* prevalence in ages 2 to 10 [*Pf*PR_2-10_]) and 561H allele frequency in Rwanda during 2014-2019 (Fig. 1; Supplemental Material 2, Table S2) – forecasts that the 561H allele currently has a national frequency of 0.24 (IQR: 0.16 – 0.34) and will reach a national frequency of 0.95 (IQR: 0.91 – 0.97) by 2033, nearly fixed as the dominant allele if current status quo conditions continue.

**Fig. 1:**
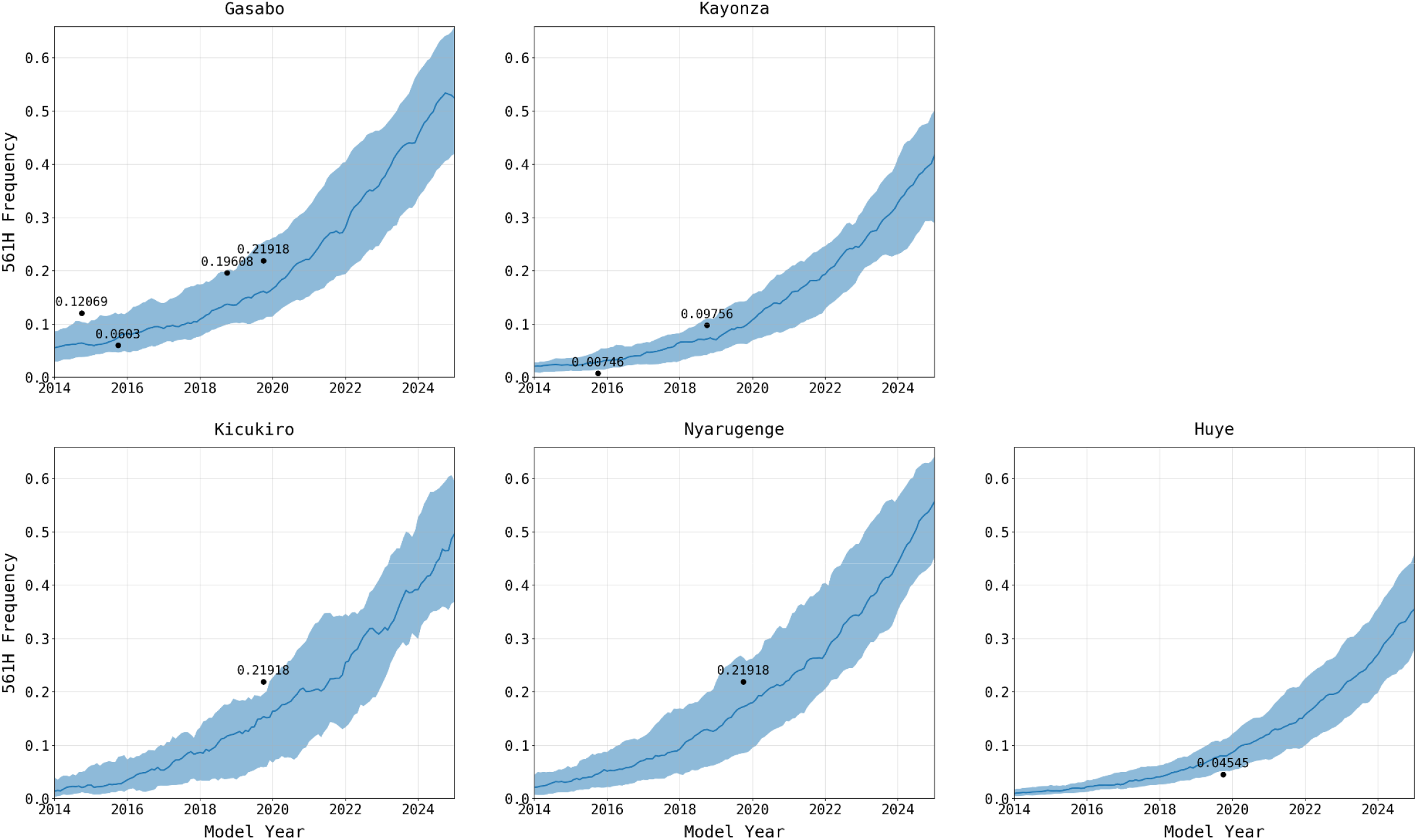
Calibration of simulated 561H allele frequency versus known frequency. 561H mutations are artificially introduced into the simulation (ten years prior to detection) in Gasabo district and allowed to evolve and increase in frequency, shown here through 2025. 561H alleles are also allowed to spread across the simulated landscape via human migration. The simulated allele frequencies in five districts (median and IQR shown with blue line and shaded area, *n* = 100) are compared to known allele-frequency data [6,7,10,11] (black dots). Gasabo, Kicukiro, and Nyarugenge districts make up Kigali City. Huye district is in southern Rwanda, and Kayonza district in in eastern Rwanda. The calibrated model is largely in agreement with known 561H spatial evolutionary patterns.

The increase in 561H allele frequency is projected to be associated with an increase in treatment failures. Based on a previously calibrated AL efficacy on commonly circulating genotypes in East and Central Africa [19], our model estimates that in 2023, presuming no interventions, 9.37% (IQR: 8.25 – 10.61) of treated *P. falciparum* malaria cases will fail treatment with AL, or about 185,000 individuals for the calendar year (median monthly average of 15,000 [IQR: 13,000 – 17,500]). This monthly average is forecast to increase to 48,000 (IQR: 44,500 – 50,000) per month by 2033. Under the current status quo conditions, the 10% treatment failure threshold recommended by the WHO for a change in first-line therapies is likely to be met or exceeded by 2025.

To evaluate the impact of potential drug policy interventions, we examined 17 national-scale drug-resistance response strategies (Table 1), with a presumed implementation date of January 1, 2023, with the objectives of *i*) reducing the near- and long-term numbers of treatment failures, and *ii*) minimizing the increase in 561H allele frequency. The interventions can broadly be placed into four categories: (1) a change in first-line recommendation to a readily available and deployable therapy, (2) a change in strategy to deployment of MFT, (3) a change to a more intensive management approach, where multiple approaches are used sequentially with different goals at different times (e.g. lowering prevalence, delaying resistance), and (4) a switch to high-efficacy triple ACTs, assuming TACTs are approved and immediately available for emergency use.

**Table 1:** Summary of the primary drug therapy interventions examined using the simulation.

| Intervention | Therapy |
| --- | --- |
| AL Extension | AL (4-day course) |
|  | AL (5-day course) |
| AL Replacement | ASAQ |
|  | DHA-PPQ |
| Multiple first-line therapies (MFT) | ASAQ (75%) + DHA-PPQ (25%) |
|  | ASAQ (50%) + DHA-PPQ (50%) |
|  | ASAQ (25%) + DHA-PPQ (75%) |
|  | AL (75%) + ASAQ (25%) |
|  | AL (50%) + ASAQ (50%) |
|  | AL (25%) + ASAQ (75%) |
|  | AL (75%) + DHA-PPQ(25%) |
|  | AL (50%) + DHA-PPQ(50%) |
|  | AL (25%) + DHA-PPQ(75%) |
| Rotation to DHA-PPQ, followed by<br>Rotation to MFT | DHA-PPQ (3 years), then AL (50%) + ASAQ (50%) |
|  | DHA-PPQ (3 years), then 5-day course of AL (50%) + ASAQ |
| Triple ACT (TACT) | ALAQ |
|  | ASMQ-PPQ |

Among alternate first-line therapies, extending the course of AL from three to four or five days is the most immediately available option due to stocks of AL already being present and available. Continued use of a 3-day AL course for five years is projected to lead to a median 561H allele frequency of 0.67 (IQR: 0.53 - 0.76), whereas 4-day AL results in a 561H allele frequency of 0.63 (IQR: 0.53 - 0.70, p = 0.0571 ; Wilcoxon Rank-Sum) and 5-day AL results in a 561H allele frequency of 0.59 (IQR: 0.48 - 0.68, p = 0.002) (Fig. 2). After five years, the national average monthly treatment failures are expected to be 27,000 (IQR: 23,000 – 30,000). These figures drop to 20,000 (IQR: 18,000 – 22,500, p < 0.0001) under 4-day AL and 16,000 (IQR: 14,000 – 18,000, p < 0.0001) under 5-day AL. Treatment failure numbers are substantially improved under a longer course of AL because of the combined effect of lower 561H frequency and higher treatment efficacy of the longer course.

**Fig. 2:**
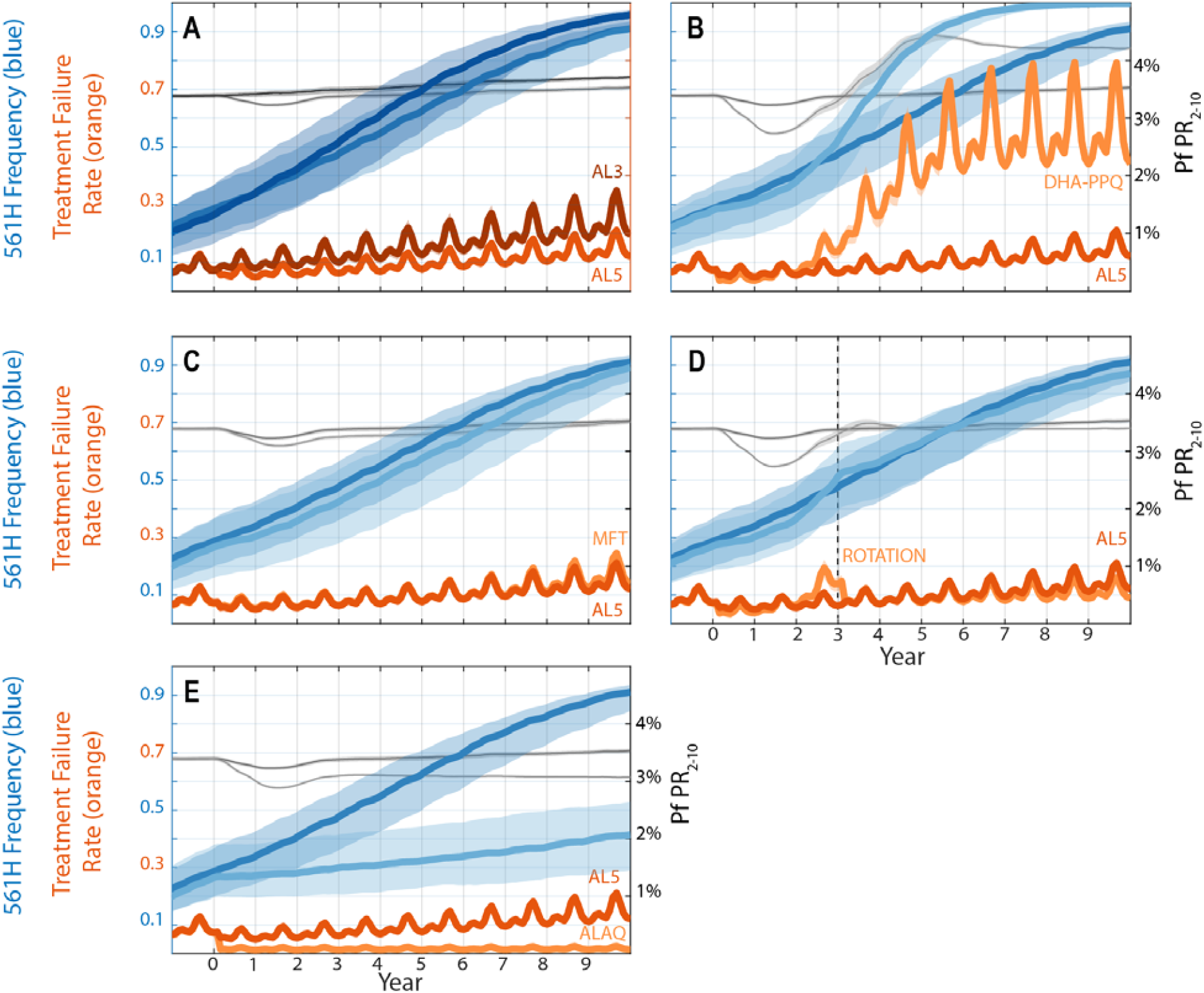
Projected 561H allele frequencies and treatment failure percentages under potential intervention scenarios. Orange lines show the percentage of treatment failures among all treated individuals in the populations; dark orange corresponding to the status quo (AL3, or three days of AL therapy), medium orange to a feasible near-term intervention (AL5, or five days of AL therapy), and light orange to other potential intervention possibilities. Blue lines show the frequency of the 561H alleles for the status quo AL3 (dark blue), feasible near-term intervention AL5 (medium blue), and other potential interventions (light blue). Black and gray lines correspond to 12-months smoothed malaria prevalence (*Pf*PR_2-10_) in status quo AL3 (black), feasible near-term intervention AL5 (medium gray), and other potential interventions (light gray). All bands show interquartile range. **(A)** Comparison between status quo (AL3, darker lines) and AL5. **(B)** Comparison between AL5 and switching to DHA-PPQ (lighter lines) as first-line therapy. Note here that under DHA-PPQ prevalence drops early (to <3%) and later rises to nearly 4.5%. This occurs because piperaquine resistance evolves quickly and reaches >0.50 genotype frequencies after three years, resulting in high prevalence and high treatment failure. **(C)** Comparison between AL5 (darker colors) and an MFT policy using ASAQ (75% of treatments) and DHA-PPQ (25%). **(D)** Comparison between AL5 (darker colors) and a rotation strategy where DHA-PPQ is used for three years, and then replaced with an MFT policy using AL5 (50%) and ASAQ (50%). **(E)** Comparison between AL5 (darker colors) and the triple therapy artemether-lumefantrine-amodiaquine (ALAQ).

Replacing AL with an alternative first-line therapy such as ASAQ or DHA-PPQ is the next easiest intervention to undertake. A switch to ASAQ gives results similar to 4-day or 5-day course of AL with a five-year 561H allele frequency of 0.60 (IQR: 0.46 - 0.67, p = 0.0029 compared to 3-day AL) and an average of 19,000 (IQR: 17,000 – 20,000, p < 0.0001) treatment failures per month within five years of deployment. However, a switch to DHA-PPQ results in an acceleration of the fixation of 561H with the allele frequency reaching 0.82 (IQR: 0.73 - 0.87, p < 0.0001) within five years, and fixation within ten years. The projected number of treatment failures is also high with a monthly average of 77,500 (IQR: 69,500 – 82,000, p < 0.0001) within five years, and 106,000 (IQR: 105,000 – 106,000, p < 0.0001) within ten years. The fixation of the 561H allele when switching to DHA-PPQ as the first-line therapy is due to the existing presence of artemisinin resistance coupled with the rapid projected evolution of piperaquine resistance leading to partner-drug failure, resulting in an evolutionary environment favorable for rapid selection for artemisinin resistance.

In contrast to extending the duration of AL treatment or replacing the first-line therapy, the introduction of MFT may pose some logistical challenges, which could be offset by the effectiveness of MFT in hindering the spread of drug-resistant genotypes (due to the more complex evolutionary environment that parasites face under MFT [20,21]). Nine combinations of AL, ASAQ, and DHA-PPQ with distribution ratios of 25/75, 50/50, and 75/25 were considered (Table 1). Barring MFTs with a high (i.e., 75%) proportion of DHA-PPQ treatments, MFTs outperformed the status quo with regards to treatment failures and 561H allele frequency (Figures 3 and 4, Supplemental Material 2, Table S1). However, only an MFT consisting of 75% ASAQ and 25% DHA-PPQ is projected to be under the 10% treatment failure threshold after five years at 9.30% (IQR: 8.28% - 10.30%, p < 0.0001 compared to 3-day AL). Within ten years, all MFT combinations are projected to exceed 10% treatment failure, although combinations of 50% AL and 50% ASAQ or 25% AL and 75% ASAQ are comparably acceptable outcomes with 14.62% (IQR: 13.74% – 15.16%, p < 0.0001) and 14.18% (IQR: 13.72% - 14.60%, p < 0.0001) treatment failure, respectively. The higher percentage of treatment failures in relation to MFTs incorporating DHA-PPQ is once again due to the loss of DHA-PPQ efficacy as piperaquine resistance evolution accelerates. After 5 years, the optimal MFT policy is projected to generate 16,000 (IQR: 14,000 – 17,000, p < 0.0001) monthly treatment failures.

**Fig. 3:**
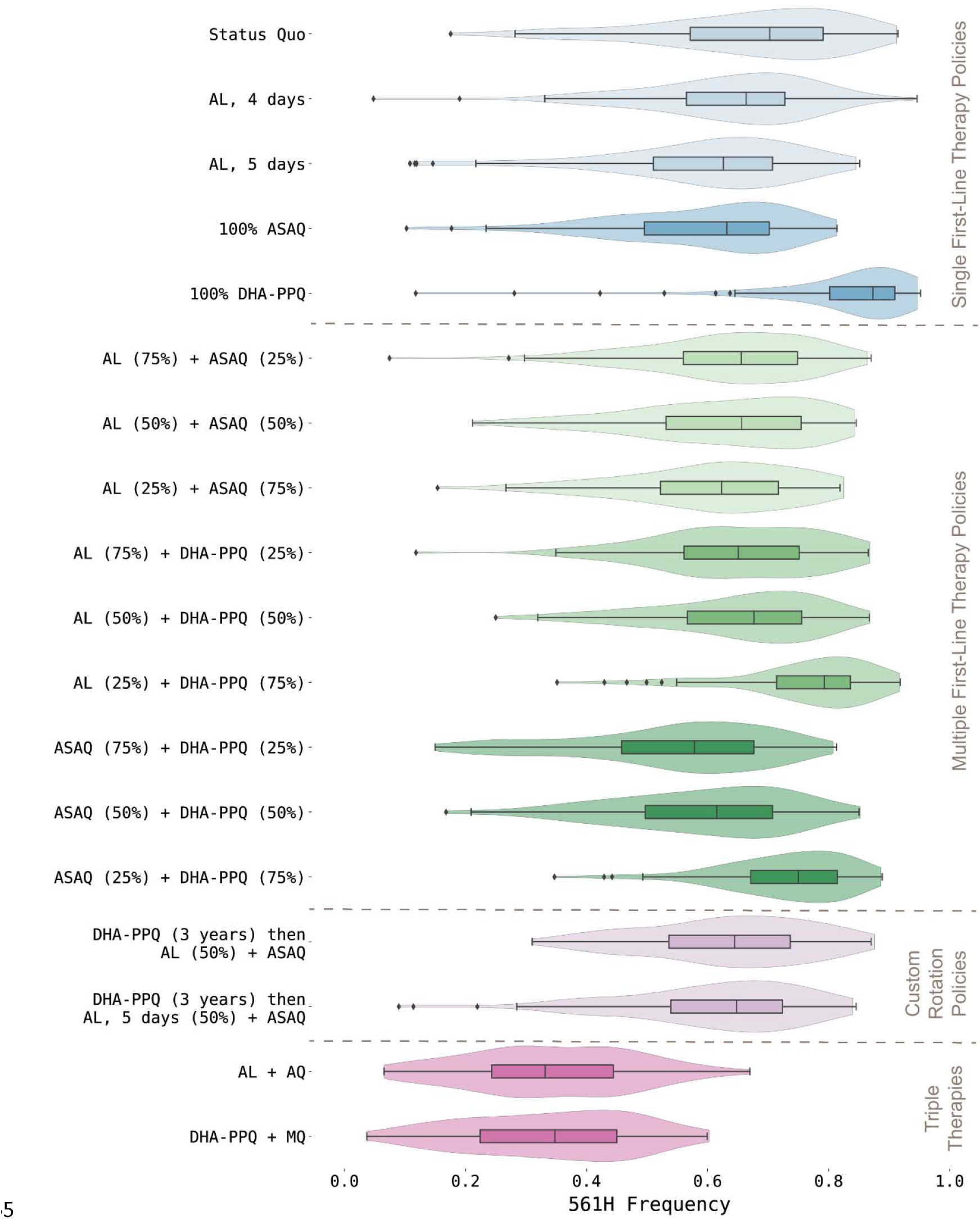
Projected 561H allele frequency after five years. Under the status quo scenario the 561H frequency is likely to be high, although this characteristic is shared by most of the other policy interventions. While the stochastic environment of the simulation allows for some outliners, the general trend is towards fixation, with complete fixation under some interventions when DHA-PPQ is used.

**Fig. 4:**
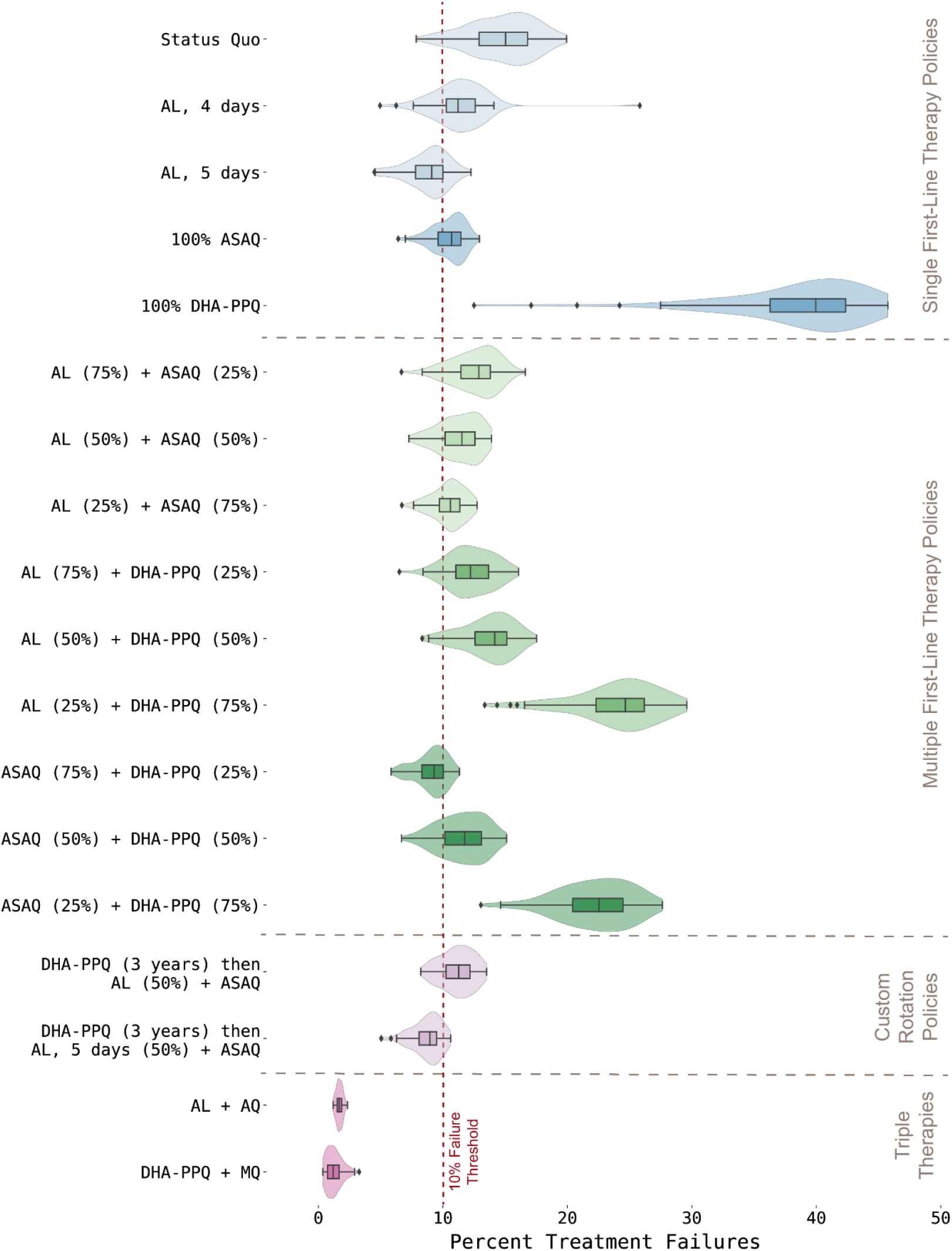
Comparison of projected treatment failures after five years. When selecting a drug policy intervention ensuring the therapeutic efficacy of the treatment is an important goal. Here we see that despite the intervention, for most interventions the treatment failure rate is likely to be above the 10% threshold recommended by the WHO for a change in policy. However, interventions involving multiple first-line therapies (MFTs) have some of the lowest treatment failure rates given the currently available therapies.

While DHA-PPQ deployment is associated with a higher frequency of 561H over the ten-year time horizon – and an associated higher level of treatment failures – over a shorter three-year time horizon DHA-PPQ is projected to be more efficacious, suggesting that a more complex rotation strategy is worth considering. To evaluate this, we introduced the use of DHA-PPQ as the first-line therapy for three years, followed by a switch to an MFT utilizing either 50% AL and 50% ASAQ or 50% 5-day AL and 50% ASAQ. As expected, this strategy performed better than the status quo in controlling the 561H allele frequency, although the percentages of treatment failures were comparable after three years. Only the strategy involving a switch to an MFT with 50% receiving a 5-day course of AL and 50% receiving ASAQ remained under the 10% threshold at the end of five years (8.95% [IQR: 8.07% - 9.50%, p = 0.0003 compared to 3-day AL]; Supplemental Material 2, Table S1) with projected monthly treatment failures of 16,000 (IQR: 14,000 – 16,500, p < 0.0001). However, the logistical complications in deploying drug rotation coupled with MFT suggest that compliance and operations would have a large influence on the success of this approach.

While the previous fifteen national-scale response strategies make use of currently available therapies, the results of past and ongoing clinical trials of the TACTs artemether-lumefantrine□and□amodiaquine (ALAQ), and artesunate-mefloquine-piperaquine□ (ASMQ-PPQ) [22], suggest that they are likely to be highly efficacious and worth considering as an emergency intervention in response to rising treatment failure rates. For these two scenarios we presume that ALAQ or ASMQ are deployed as first-line therapy, replacing AL. As expected based upon previous modeling studies [23,24], in our analysis TACTs outperformed all other drug policy interventions with ALAQ resulting in average monthly treatment failures of 2,800 (IQR: 2,500 – 3,000, p < 0.0001 compared to 3-day AL) and a 561H allele frequency of 0.33 (IQR: 0.24 – 0.44, p < 0.0001) after five years, while ASMQ-PPQ resulted in 1,800 (IQR: 1,000 – 2,500, p < 0.0001) monthly treatment failures and a 561H allele frequency of 0.33 (IQR: 0.22 - 0.43, p < 0.0001). However, the usage of ASMQ-PPQ comes an increased risk of DHA-PPQ failure given projected treatment failures of 16.48% (IQR: 11.34% - 19.38%, p < 0.0001) within ten years of deployment. In contrast, treatment failures are likely to still be low if ALAQ is deployed reaching only 1.82% (IQR: 1.65% – 2.02%, p < 0.0001) within ten years of deployment.

These modeled scenarios assume that all courses of treatments will be completed in full; however, in practice 100% compliance is unlikely and adherence rates may be complicated by under-dosing, over-dosing, and formulation design (i.e., fixed dose combination vs. co-packaged blister packs), resulting in real-world compliance rates between 65% and 90% for a three-day course of treatment [25,26]. To evaluate the possible impacts that compliance with treatment regimens would have, additional scenarios were evaluated in which ASAQ, DHA-PPQ, and 3-, 4-, or 5-day courses of AL were administered using low (25% to 70%), moderate (50% to 80%), and high (70% to 90%) compliance rates for complete courses (Supplemental Material 2, Table S4). As expected, failure to comply with the prescribed course of treatment results in an increase in treatment failures; however, extended courses of AL (four or five days) along with ASAQ still outperform perfect compliance with a 3-day course of AL (Supplemental Material 2, Table S3). No differences were seen in policy prioritization when evaluating scenarios with imperfect compliance.

## Discussion

The projected national frequency of the 561H allele in Rwanda under a continuation of status quo treatment with a 3-day course of artemether-lumefantrine indicates that treatment failures will increase over the next ten years and drug policy interventions are required to mitigate this risk as much as possible. The current spread of the 561H allele to other districts from its initial apparent emergence in Gasabo, together with confirmation of 561H [6,7,10,11] at frequencies similar to those projected in our simulation, suggests that the 561H allele is likely present throughout Rwanda.

Our findings suggest that over a five-year time horizon extending the use of AL from three to five days would hold treatment failures under 10% (projected median 9.11% [IQR: 7.79% - 10.00%]), and switching to an MFT strategy is likewise worth considering, with the optimal MFT approach – a 75% ASAQ and 25% DHA-PPQ deployment – projected to lower treat failures to a median 9.30% [IQR: 8.28% - 10.03%] after five years. Extending the course of treatment with AL has minimal logistical considerations beyond ensuring sufficient quality of doses being distributed and appears to be beneficial compared to the status quo scenario. While there are concerns regarding the cardiotoxicity of antimalarial drugs [27,28], the incidence of adverse cardiac events recorded during clinical trials has been low [29]. Switching to one of several MFT options where future treatment failure rates can be kept close to 10% will ensure there are no concerns with extended artemisinin dosing, and the success of these MFT deployments will depend on operational capability around the supply and distribution of ACTs. Based on the structure of the distribution network for antimalarials in Rwanda and the current availability of different ACTs, an MFT deployment is more likely to be feasible than a custom rotation approach.

These findings must be contextualized against the limitations that are inherent in modeling studies. First, due to the introduction of AL in 2006, other drug-resistance mutations – such as those associated with lumefantrine resistance – are likely to be present in Rwanda. The enabling of mutations in model year 2014 represents a ‘model fitting’ compromise made necessary due to the stochastic nature of the rare mutation emergence process [19,30] from 2006 to 2014 that has not been systematically captured in any known data collections. Higher (or lower) frequencies of partner-drug sensitivity would affect the selection for the 561H mutant. Furthermore, while the model calibration may suggest that 561H initially appeared in Gasabo district around 2004, the design of the simulation renders it incapable of fitting the highly stochastic process that would have driven the initial appearance and spread of 561H.

Second, pyronaridine-artesunate deployment was not considered in our simulations as pyronaridine-resistant falciparum phenotypes have not yet been described. Assuming similar rates of emergence and similar drops in ACT efficacy due to future pyronaridine resistance, pyronaridine-artesunate would likely make a positive contribution as an addition to any of the MFT strategies shown in Table 1. Finally, due to our model calibration being centered on 2017 estimates of falciparum prevalence, our current simulation set-up does not capture the gains made in malaria control in Rwanda during 2017-2021. Annual falciparum cases have been reported to be around one million for the 2021-2022 fiscal period (July to June) for an annual incidence of 76/1000. If these gains hold for the remainder of the decade, the absolute number of monthly treatment failure counts reported in our analysis could be lower by a factor of two or three. The relative benefits of the different strategies should stay the same, however this would need to be confirmed with the latest case counts and drug coverage patterns.

Crucially, current National Malaria Control Program activities to expand indoor residual spraying activities (from 2-8 districts last decade to 12-15 districts this decade) and to introduce synergistic insecticide impregnated nets (piloted in 2019-2020 with distribution planned for 2023 [31]) are likely to reduce biting rates and case numbers in the coming years. A recent pilot study in rural part of Gasabo district with high malaria risk showed that larviciding activity had a moderate effect on reducing incidence; NMCP also expects to expand larviciding as part of a new set of malaria control activities. These interventions may prove beneficial in the fight against drug resistance as they may eliminate pockets of transmission, including drug-resistant alleles, and are likely to slow the overall geographic spread of all genotypes.

Without additional interventions targeting drug resistance and general malaria transmission, the results of this modeling study underscore what has already been demonstrated: drug resistance can spread rapidly once it is established, and this could leave National Malaria Control Programs with limited forewarning to deploy responses. The identification of 561H in Uganda [32] suggests that the spread of the 561H mutant is currently in progress, and this spread is likely to co-occur with that of other markers for artemisinin resistance within sub-Saharan Africa [32–34].

It is important to highlight that for the majority of national drug-policy intervention scenarios considered here, treatment failures are projected to exceed 10% within five years and may reach as high as 40% if DHA-PPQ were to be deployed as the sole first-line therapy without rotation (Supplemental Material 2, Table S1). As such, emphasis should be placed upon the development of next generation therapies along with continued evaluation of therapies such pyronaridine-artesunate, ASMQ, and extended courses of ACT. According to model projections, the most effective among these choices is adoption of TACTs following completion of clinical trials and regulatory approval which is projected to slow down the spread of 561H and greatly reduce treatment failures. Early introduction of TACTs, before crossing the WHO first-line therapy treatment failure threshold of 10%, would need to be accompanied by appropriate public health communication and an assessment of acceptability in affected communities [35]. Ultimately, the results of the use of TACTs within the context of this study are promising and suggests that if approved, they could prove to be highly effective as a bridge to next generation malaria therapies.

## Methods

A previously validated spatial, stochastic, individually-based model was used as the basis for the study [36,37], and a new model calibration was conducted to match the malaria prevalence of the 30 administrative districts of Rwanda. The Malaria Atlas Project mean *Pf*PR_2-10_ projections for 2017 [38] were used as the basis to calibrate local transmission parameters on a 5-by-5 km (25 sq.km) scale in Rwanda (Supplemental Material 1, §1.1). The seasonal variation in malaria transmission was coupled to seasonal rainfall (Supplemental Material 1, §1.2), consistent with the general coupling of seasonal transmission and increased rainfall. Treatment seeking and treatment coverage data were obtained from the 2019-20 Demographic and Health Survey [39], with treatment coverage ranging from 53.3% to 71.8% across districts (Supplemental Material 1, §1.3). Under the baseline model – or status quo –conditions, all treated individuals in the model receive a 3-day course of AL, with full compliance with the course of treatment. No private market drugs are incorporated, consistent with survey results [39]. During model execution, individuals move around the simulated landscape in a manner consistent with previous travel studies for SSA [40,41], and carry any *P. falciparum* clones to these destinations. The carried clones may then enter into circulation within a new region if the traveling individual is bitten by a mosquito, which then proceeds to infect another individual.

With these prevalence and treatment calibrations, the model produces a symptomatic malaria incidence ranging from 9.7 per 1000 (Burera district) to 452.53 per 1000 (Gisagara district) annual cases. This sums to about 217 annual cases per 1000 on a national level, consistent with reporting that the incidence rate has been declining from a high of 403 per 1000 in 2016 [42] (Supplemental Material 1, §3, §4). Despite concerns that the global COVID-19 pandemic would impact malaria control measures, a mixed-methods study of three high-endemic districts (Gasabo, Kayonza, and Rwamagana) suggests that while the distribution of malaria testing shifted, the overall decline in uncomplicated malaria cases has continued through 2020 [43]. This indicates that the model calibration may be overestimating case numbers in 2020 and later.

After calibration of prevalence, incidence, and treatment coverage, the simulation’s genotype frequency and trajectory were matched to field observations of 561H allele frequencies. Seven data points on *pfkelch* 561H mutation are available at district-level: four close to the capital Kigali [6,7,10], two from the eastern district of Kayonza [6,7], and one from the southern district of Huye [11]. To calibrate to these values the simulation was seeded with a single mutation of 561H genotypes in Gasabo district (Kigali province) to generate a slowly growing exponential curve with district frequencies that are consistent with measured values (Fig. **1**). This results in a mean national frequency ranging from 0.01 in 2014 to 0.12 in 2020 (see Supplemental Material 1, Figure S6). In the event that the seeded 561H genotypes went extinct before 2014, the replicate was discarded from analysis.

Since the mutation rate for 561H is not known, these artificially introduced 561H genotypes are the only means by which 561H can be introduced into the simulation. The spatial spread of 561H is driven by human movement and migration within the simulation. To account for the ability of ACT partner-drug resistance to accelerate the fixation of artemisinin resistance [19], mutations affecting other alleles (e.g., *pfcrt, pfmdr1*, etc.) are enabled in the simulation on January 1, 2014 following the completion of model burn-in using a previously calibrated value [19]. This produces a slight model delay in *pfcrt* and *pfmdr1* mutations that are associated with the use of lumefantrine given the adoption of AL by Rwanda in 2006, likely resulting in a slight model bias against lumefantrine resistance.

As a common point of comparison, a baseline scenario (also called “status quo”) was run (*n* = 100 replicates) in which the calibrated model was simulated for a ten-year window past the point of intervention (January 1, 2023, to January 1, 2033). Seventeen drug policy intervention scenarios (*n* = 100 replicates each) were evaluated within the simulation (see Table 1), and all interventions are implemented at the same time with no delay between the introduction and the change in therapies received by individuals. Four of the evaluated interventions were for a change in first-line therapy to a 4-day course of AL, 5-day course of AL, ASAQ, or DHA-PPQ. Nine of the interventions were 2-therapy MFT approaches with drug distribution proportions ranging from 25/75 to 50/50 to 75/25 (see Table 1). Two interventions considered short-term DHA-PPQ use first (for three years) followed by replacement of DHA-PPQ with one of two MFT strategies using AL and ASAQ (Table 1). Finally, two interventions considered replacement of AL with one of two triple therapies (ALAQ and ASMQ-PPQ) which are yet to be approved [22,35]. Adherence to drug course regimen was evaluated to determine if patient compliance with three-to-five-day drug courses had major effects on strategy choice. The five-year endpoints are presented in the main text, with three- and ten-year endpoints referred to as needed; comprehensive results for all endpoints can be found in Supplemental Material 2, Tables S1 and S3.

Differences between groups of simulation results (i.e., comparing 100 simulations each from two scenarios) are tested with non-parametric Wilcoxon rank-sum tests and all p-values lower than 10^−4^ are shown as 10^−4^.

## Data Availability

All data and source code used in the present work is available online at https://github.com/bonilab/malariaibm-spatial-Rwanda-561H

## References

1. Ariey F, Witkowski B, Amaratunga C, Beghain J, Langlois A-C, Khim N, et al. A molecular marker of artemisinin-resistant Plasmodium falciparum malaria. Nature. 2014;505:50–5.

2. Ashley EA, Dhorda M, Fairhurst RM, Amaratunga C, Lim P, Suon S, et al. Spread of Artemisinin Resistance in Plasmodium falciparum Malaria. N Engl J Med. Massachusetts Medical Society; 2014;371:411–23.

3. World Health Organization. Report on antimalarial drug efficacy, resistance and response: 10 years of surveillance (2010 - 2019) [Internet]. Geneva: World Health Organization; 2020 p. 78. Available from: https://apps.who.int/iris/bitstream/handle/10665/336692/9789240012813-eng.pdf

4. World Health Organization. World Malaria Report 2021 [Internet]. Geneva, Switzerland: World Health Organization; 2021 [cited 2022 Jun 17]. Available from: https://www.who.int/teams/global-malaria-programme/reports/world-malaria-report-2021

5. Fidock DA, Rosenthal PJ. Artemisinin resistance in Africa: How urgent is the threat? Med. 2021;2:1287–8.

6. Uwimana A, Legrand E, Stokes BH, Ndikumana J-LM, Warsame M, Umulisa N, et al. Emergence and clonal expansion of in vitro artemisinin-resistant Plasmodium falciparum kelch13 R561H mutant parasites in Rwanda. Nature Medicine [Internet]. 2020; Available from: https://doi.org/10.1038/s41591-020-1005-2

7. Uwimana A, Umulisa N, Venkatesan M, Svigel SS, Zhou Z, Munyaneza T, et al. Association of Plasmodium falciparum kelch13 R561H genotypes with delayed parasite clearance in Rwanda: an open-label, single-arm, multicentre, therapeutic efficacy study. The Lancet Infectious Diseases. 2021;21:1120–8.

8. Karema C, Aregawi MW, Rukundo A, Kabayiza A, Mulindahabi M, Fall IS, et al. Trends in malaria cases, hospital admissions and deaths following scale-up of anti-malarial interventions, 2000–2010, Rwanda. Malaria Journal. 2012;11:236.

9. Amaratunga C, Andrianaranjaka VH, Ashley E, Bethell D, Björkman A, Bonnington CA, et al. Association of mutations in the Plasmodium falciparum Kelch13 gene (Pf3D7_1343700) with parasite clearance rates after artemisinin-based treatments—a WWARN individual patient data meta-analysis. BMC Medicine. 2019;17:1.

10. Straimer J, Gandhi P, Renner KC, Schmitt EK. High Prevalence of Plasmodium falciparum K13 Mutations in Rwanda Is Associated With Slow Parasite Clearance After Treatment With Artemether-Lumefantrine. The Journal of Infectious Diseases [Internet]. 2021 [cited 2021 Sep 7]; Available from: https://doi.org/10.1093/infdis/jiab352

11. Bergmann C, van Loon W, Habarugira F, Tacoli C, Jäger J, Savelsberg D, et al. Increase in Kelch 13 Polymorphisms in Plasmodium falciparum, Southern Rwanda. Emerging Infectious Disease journal. 2021;27:294.

12. Kirby R, Giesbrecht D, Karema C, Watson O, Lewis S, Munyaneza T, et al. Examining the early distribution of the artemisinin-resistant Plasmodium falciparum kelch13 R561H mutation in Rwanda. bioRxiv. 2022;2022.10.26.513523.

13. WHO Global Malaria Programme. WHO Guidelines for Malaria - 25 November 2022 [Internet]. World Health Organization; 2022 Nov p. 425. Report No.: WHO/UCN/GMP/2022.01 Rev.3. Available from: https://app.magicapp.org/#/guideline/6810

14. World Health Organization. Seasonal Malaria Chemoprevention with Sulfadoxine-Pyrmethamine Plus Amodiaquine in Children: A Field Guide [Internet]. Geneva, Switzerland: World Health Organization; 2013 [cited 2022 Jul 18]. Available from: https://apps.who.int/iris/bitstream/handle/10665/85726/9789241504737_eng.pdf

15. Whegang Youdom S, Tahar R, Basco LK. Comparison of anti-malarial drug efficacy in the treatment of uncomplicated malaria in African children and adults using network meta-analysis. Malaria Journal. 2017;16:311.

16. van Vugt M, Looareesuwan S, Wilairatana P, McGready R, Villegas L, Gathmann I, et al. Artemether-lumefantrine for the treatment of multidrug-resistant falciparum malaria. Transactions of The Royal Society of Tropical Medicine and Hygiene. 2000;94:545–8.

17. Tun KM, Jeeyapant A, Myint AH, Kyaw ZT, Dhorda M, Mukaka M, et al. Effectiveness and safety of 3 and 5 day courses of artemether–lumefantrine for the treatment of uncomplicated falciparum malaria in an area of emerging artemisinin resistance in Myanmar. Malaria Journal. 2018;17:258.

18. Onyamboko Marie A., Hoglund Richard M., Lee Sue J., Kabedi Charlie, Kayembe Daddy, Badjanga Benjamin B., et al. A Randomized Controlled Trial of Three-versus Five-Day Artemether-Lumefantrine Regimens for Treatment of Uncomplicated Plasmodium falciparum Malaria in Pregnancy in Africa. Antimicrobial Agents and Chemotherapy. American Society for Microbiology; 2020;64:e01140–19.

19. Watson OJ, Gao B, Nguyen TD, Tran TN-A, Penny MA, Smith DL, et al. Pre-existing partner-drug resistance to artemisinin combination therapies facilitates the emergence and spread of artemisinin resistance: a consensus modelling study. The Lancet Microbe [Internet]. 2022; Available from: https://www.sciencedirect.com/science/article/pii/S2666524722001550

20. Boni MF, Smith DL, Laxminarayan R. Benefits of using multiple first-line therapies against malaria. Proc Natl Acad Sci USA. 2008;105:14216.

21. Boni MF, White NJ, Baird JK. The Community As the Patient in Malaria-Endemic Areas: Preempting Drug Resistance with Multiple First-Line Therapies. PLOS Medicine. Public Library of Science; 2016;13:e1001984.

22. van der Pluijm RW, Tripura R, Hoglund RM, Pyae Phyo A, Lek D, ul Islam A, et al. Triple artemisinin-based combination therapies versus artemisinin-based combination therapies for uncomplicated Plasmodium falciparum malaria: a multicentre, open-label, randomised clinical trial. The Lancet. 2020;395:1345–60.

23. Kunkel A, White M, Piola P. Novel anti-malarial drug strategies to prevent artemisinin partner drug resistance: A model-based analysis. PLOS Computational Biology. Public Library of Science; 2021;17:e1008850.

24. Nguyen TD, Gao B, Amaratunga C, Dhorda M, Tran TN-A, White NJ, et al. Preventing antimalarial drug resistance with triple artemisinin-based combination therapies. medRxiv. 2022;2022.10.21.22281347.

25. Cohen JL, Yavuz E, Morris A, Arkedis J, Sabot O. Do patients adhere to over-the-counter artemisinin combination therapy for malaria? evidence from an intervention study in Uganda. Malaria Journal. 2012;11:83.

26. Banek K, Webb EL, Smith SJ, Chandramohan D, Staedke SG. Adherence to treatment with artemether–lumefantrine or amodiaquine–artesunate for uncomplicated malaria in children in Sierra Leone: a randomized trial. Malaria Journal. 2018;17:222.

27. White NJ. Cardiotoxicity of antimalarial drugs. The Lancet Infectious Diseases. 2007;7:549–58.

28. Chan XHS, Haeusler IL, Win YN, Pike J, Hanboonkunupakarn B, Hanafiah M, et al. The cardiovascular effects of amodiaquine and structurally related antimalarials: An individual patient data meta-analysis. PLOS Medicine. Public Library of Science; 2021;18:e1003766.

29. Haeusler IL, Chan XHS, Guérin PJ, White NJ. The arrhythmogenic cardiotoxicity of the quinoline and structurally related antimalarial drugs: a systematic review. BMC Medicine. 2018;16:200.

30. Pongtavornpinyo W, Hastings IM, Dondorp A, White LJ, Maude RJ, Saralamba S, et al. Probability of emergence of antimalarial resistance in different stages of the parasite life cycle. Evol Appl. John Wiley & Sons, Ltd; 2009;2:52–61.

31. Gansané A, Candrinho B, Mbituyumuremyi A, Uhomoibhi P, NFalé S, Mohammed AB, et al. Design and methods for a quasi-experimental pilot study to evaluate the impact of dual active ingredient insecticide-treated nets on malaria burden in five regions in sub-Saharan Africa. Malaria Journal. 2022;21:19.

32. Asua V, Conrad MD, Aydemir O, Duvalsaint M, Legac J, Duarte E, et al. Changing Prevalence of Potential Mediators of Aminoquinoline, Antifolate, and Artemisinin Resistance Across Uganda. The Journal of Infectious Diseases. 2021;223:985–94.

33. Asua V, Vinden J, Conrad MD, Legac J, Kigozi SP, Kamya MR, et al. Changing Molecular Markers of Antimalarial Drug Sensitivity across Uganda. Antimicrobial Agents and Chemotherapy. 2019;63.

34. Balikagala B, Fukuda N, Ikeda M, Katuro OT, Tachibana S-I, Yamauchi M, et al. Evidence of Artemisinin-Resistant Malaria in Africa. N Engl J Med. Massachusetts Medical Society; 2021;385:1163–71.

35. Tindana P, de Haan F, Amaratunga C, Dhorda M, van der Pluijm RW, Dondorp AM, et al. Deploying triple artemisinin-based combination therapy (TACT) for malaria treatment in Africa: ethical and practical considerations. Malaria Journal. 2021;20:119.

36. Nguyen TD, Olliaro P, Dondorp AM, Baird JK, Lam HM, Farrar J, et al. Optimum population-level use of artemisinin combination therapies: a modelling study. Lancet Glob Health. Elsevier; 2015;3:e758–66.

37. Zupko RJ, Nguyen TD, Somé AF, Tran TN-A, Gerardin J, Dudas P, et al. Long-term effects of increased adoption of artemisinin combination therapies in Burkina Faso. PLOS Global Public Health. Public Library of Science; 2022;2:e0000111.

38. Weiss DJ, Lucas TCD, Nguyen M, Nandi AK, Bisanzio D, Battle KE, et al. Mapping the global prevalence, incidence, and mortality of Plasmodium falciparum, 2000–17: a spatial and temporal modelling study. Lancet. Elsevier; 2019;394:322–31.

39. National Institute of Statistics of Rwanda, Ministry of Health, The DHS Program. Rwanda Demographic and Health Survey 2019-20: Key Indicators [Internet]. Kigali, Rwanda, and Rockville, Maryland, USA: National Institute of Statistics of Rwanda; 2020 Oct p. 53. Available from: https://www.statistics.gov.rw/publication/demographic-and-health-survey-20192020-key-indicators

40. Wesolowski A, Stresman G, Eagle N, Stevenson J, Owaga C, Marube E, et al. Quantifying travel behavior for infectious disease research: a comparison of data from surveys and mobile phones. Scientific Reports. 2014;4:5678.

41. Wesolowski A, O’Meara WP, Eagle N, Tatem AJ, Buckee CO. Evaluating Spatial Interaction Models for Regional Mobility in Sub-Saharan Africa. PLoS Comput Biol. 2015;11:e1004267.

42. U.S. President’s Malaria Initiative. U.S. President’s Malaria Initiative Rwanda: Malaria Operational Plan FY 2020 [Internet]. Available from: https://www.pmi.gov/fy-2020-rwanda-malaria-operational-plan/

43. Hakizimana D, Ntizimira C, Mbituyumuremyi A, Hakizimana E, Mahmoud H, Birindabagabo P, et al. The impact of Covid-19 on malaria services in three high endemic districts in Rwanda: a mixed-method study. Malaria Journal. 2022;21:48.

