## Supplemental Material 1 for "Potential policy interventions for slowing the spread of artemisinin-resistant *pfkelch* R561H mutations in Rwanda"

**Supplemental Material for “Optimal drug policy interventions for slowing the spread of the *pfkelch* R561H artemisinin-resistance mutation in Rwanda”, Zupko et al. (2022)**

### 1 Model Spatial Data

#### 1.1 Overview

Figure S1. Projected annual mean *Pf*PR_2-10_ by Malaria Atlas Project (version 2019) for Rwanda (Weiss et al., 2019). Map prepared by the authors using ArcGIS Pro (version 3.0) using administrative brounderies from World Bank Group (2018, 2020), geographic data from Rwanda National Institute of Statistics (2018), and prevalence data from the Malaria Atlas Project (Weiss et al., 2019).

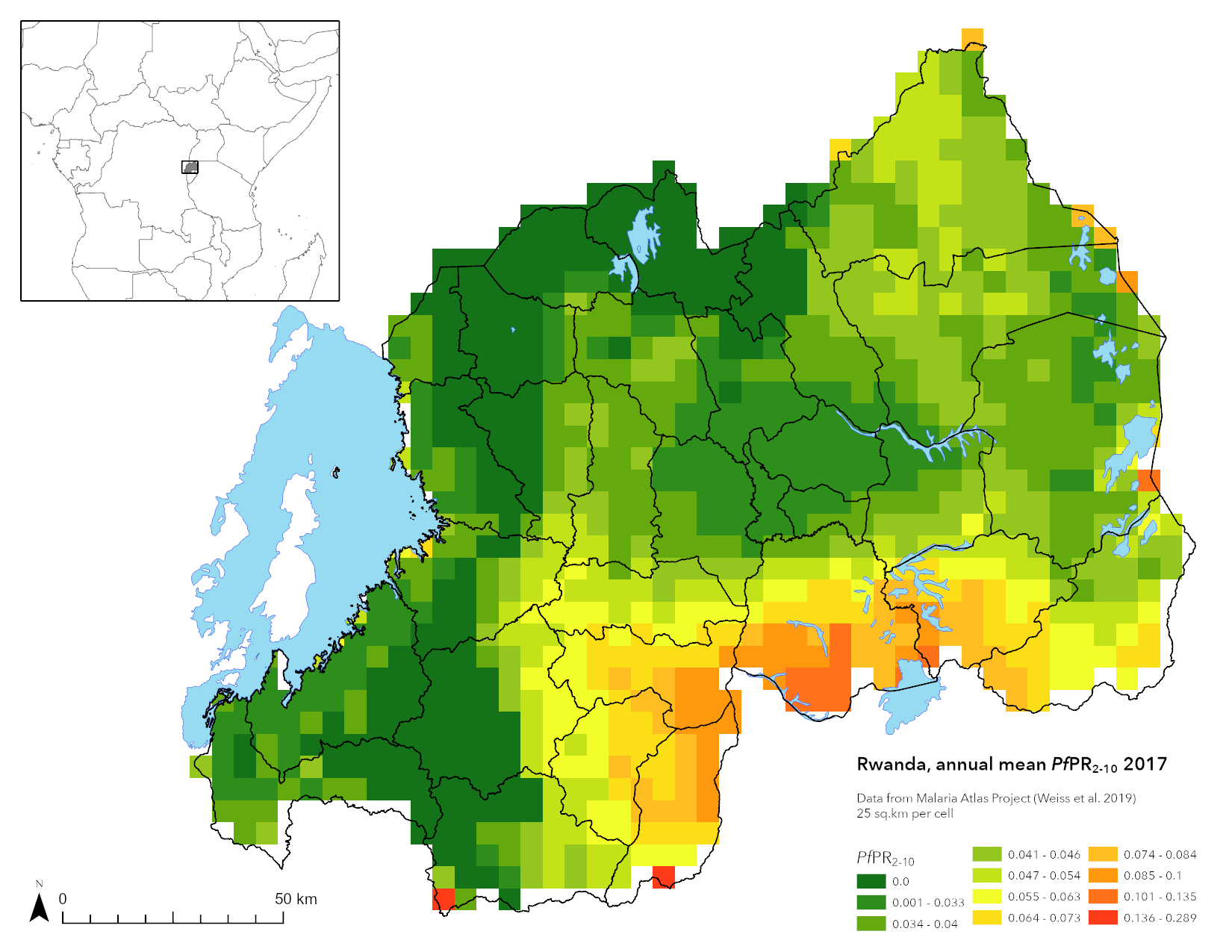

The primary spatial unit in the simulation is the cell, which represents a 5km-by-5km (25 sq.km) location, with the size selected due to the Malaria Atlas Project *Pf*PR_2-10_ projections using the same footprint (Weiss et al., 2019). This results in 979 cells being used to cover 24,475 sq.km or about 93% of the total area of Rwanda (Figure S1). Cells that are predominately water are not simulated, and some discrepancy in area is due to clipping along national borders. During model initialization, spatial data in the form of national districts, population (Section 2), access to treatment (Section 2), and the beta (i.e., transmission parameter) for each cell is loaded (Section 3). A single seasonality calibration (Section 1.2) is used for all cells in the simulation. During model execution the results are aggregated from the cell up to the district and national level as needed.

#### 1.2 Seasonality

Figure S2*.* Comparison of the ten-year daily average rainfall (Top) with the calibrated adjustment in the beta used in the simulation (Bottom).

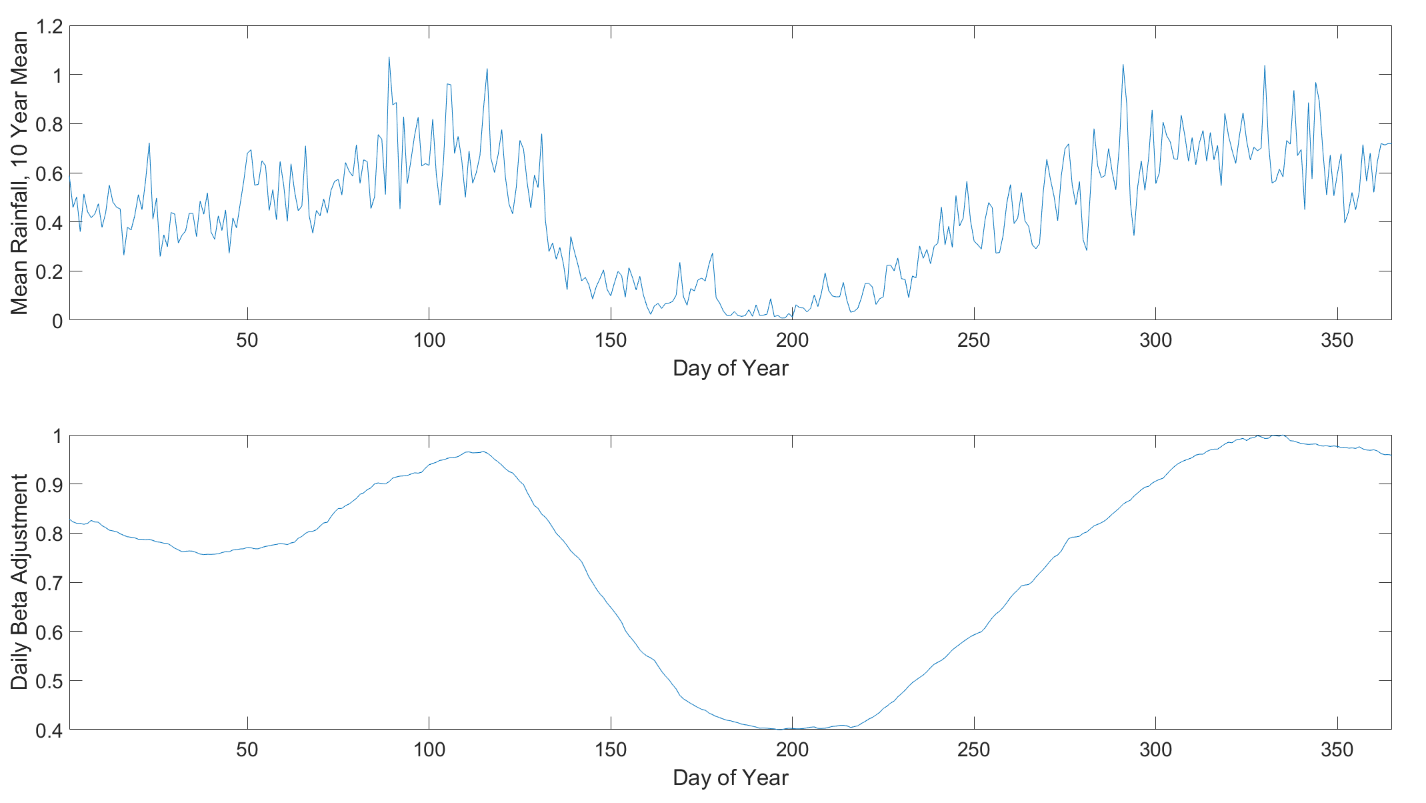

The pattern of seasonal transmission used within the simulation is based upon the correlation between rainfall and the *Anopheles* mosquito and presumes that transmission will begin to increase as favorable conditions increase (Figure S2). Thus, the beta is adjusted up – or down – using an adjustment value that calculated by first finding the ten-year daily average rainfall for Rwanda, based upon data from the data from ERA5 global climate and weather projections (Copernicus Climate Change Service 2017). The data was clipped to the boarders of Rwanda using Google Earth Engine for January 2009 to December 2019, inclusive, and then smoothed and shifted by ten days using a MATLAB script.^[1]^ With the ten-day offset was selected on the basis *Anopheles gambiae* lifecycle. The calculated adjustment is then imported during model initialization and applied during execution.

### 2 Demographics, Mortality, and Treatment Seeking

Table S1. Population distribution of Rwanda (Malaria and Other Parasitic Diseases Division of the Rwanda Biomedical Center Ministry of Health [Rwanda] & ICF, 2018)

| **Age Band** | **Population** | **Age Band** | **Population** |
| --- | --- | --- | --- |
| < 5 | 15.1 | 45 – 49 | 3.6 |
| 5 – 9 | 14.1 | 50 – 54 | 3 |
| 10 – 14 | 14.5 | 55 – 59 | 3.1 |
| 15 – 19 | 9.6 | 60 – 64 | 2.2 |
| 20 – 24 | 7.3 | 65 – 69 | 1.2 |
| 25 – 29 | 6.9 | 70 – 74 | 0.9 |
| 30 – 34 | 6.5 | 75 – 79 | 0.6 |
| 35 – 39 | 6.1 | 80+ | 0.8 |
| 40 – 44 | 4.5 |  |  |

The estimated population of Rwanda was 12,663,116 in 2020 with a crude birth rate of 28.8 per 1,000 (National Institute of Statistics of Rwanda, 2021). The population skews younger with about 54.3% being under the age of 20 (Table S1). Due to malaria continue to have a significant impact upon the mortality rates within Rwanda (Table S2), the mortality rate applied to the population was adjusted to remove the deaths that were attributed to malaria (Table S3) with individuals that reach the age of 100 being removed from the simulation. When individuals are infected with the parasite, upon exhibiting clinical symptoms, they seek treatment based upon the surveyed treatment seeking behavior for the provenience that contains the cell they are currently in (Table S4) with no distinction between under-5 and over-5 treatment seeking rates.

Table S2. Proportion of deaths that are attributable to malaria for 2012 to 2018 (Republic of Rwanda, Ministry of Health, n.d., p. 82)

|  | **2012** | **2013** | **2014** | **2015** | **2016** | **2017** | **2018** |
| --- | --- | --- | --- | --- | --- | --- | --- |
| < 5 due to Malaria | 48 | 64 | 173 | 130 | 158 | 139 | 84 |
| < 5 all deaths | 1647 | 1854 | 5080 | 4385 | 4162 | 3842 | 3802 |
| > 5 due to Malaria | 314 | 401 | 416 | 401 | 685 | 244 | 241 |
| > 5 deaths | 7379 | 7275 | 4356 | 7175 | 7846 | 5464 | 6249 |
| < 5 malaria proportional | 0.029 | 0.035 | 0.034 | 0.03 | 0.038 | 0.036 | 0.022 |
| > 5 malaria proportional | 0.043 | 0.055 | 0.096 | 0.056 | 0.087 | 0.045 | 0.039 |

Table S3. Malaria adjusted mortality rate used in the simulation, derived from UN population projections (UN, 2019).

| **Age Band** | **Mortality Rate** |
| --- | --- |
| 0 – 1 | 0.02641 |
| 2 | 0.00202 |
| 3 | 0.00202 |
| 4 | 0.00202 |
| 5 | 0.00198 |
| 6 | 0.00247 |
| 7 | 0.00247 |
| 8 | 0.00247 |
| 9 | 0.00247 |
| 10 | 0.00247 |
| 11 | 0.00247 |
| 12 – 15 | 0.00247 |
| 16 – 20 | 0.00455 |
| 21 – 60 | 0.00455 |
| 61 – 100 | 0.05348 |

Table S4. Treatment seeking behavior in Rwanda (National Institute of Statistics of Rwanda et al., 2020).

| **Province** | **Treatment Seeking** |
| --- | --- |
| Northern Province | 53.3% |
| Eastern Province | 63.4% |
| Kigali City | 71.8% |
| Southern Province | 61.6% |
| Western Province | 63.3% |

### 3 Model Calibration and Validation

#### 3.1 P. falciparum malaria cases and prevalence

Figure S3. (left) Comparison of simulated *Pf*PR_2-10_ versus the reference *Pf*PR_2-10_ from the Malaria Atlas Project (Weiss et al., 2019), note that simulated results are typically with ±10% of the reference value, although low *Pf*PR_2-10_ values tend to be higher. (right) The total simulated clinical cases (dark gray) versus the reported clinical cases (light gray). As expected, there are fewer reported cases than total cases.

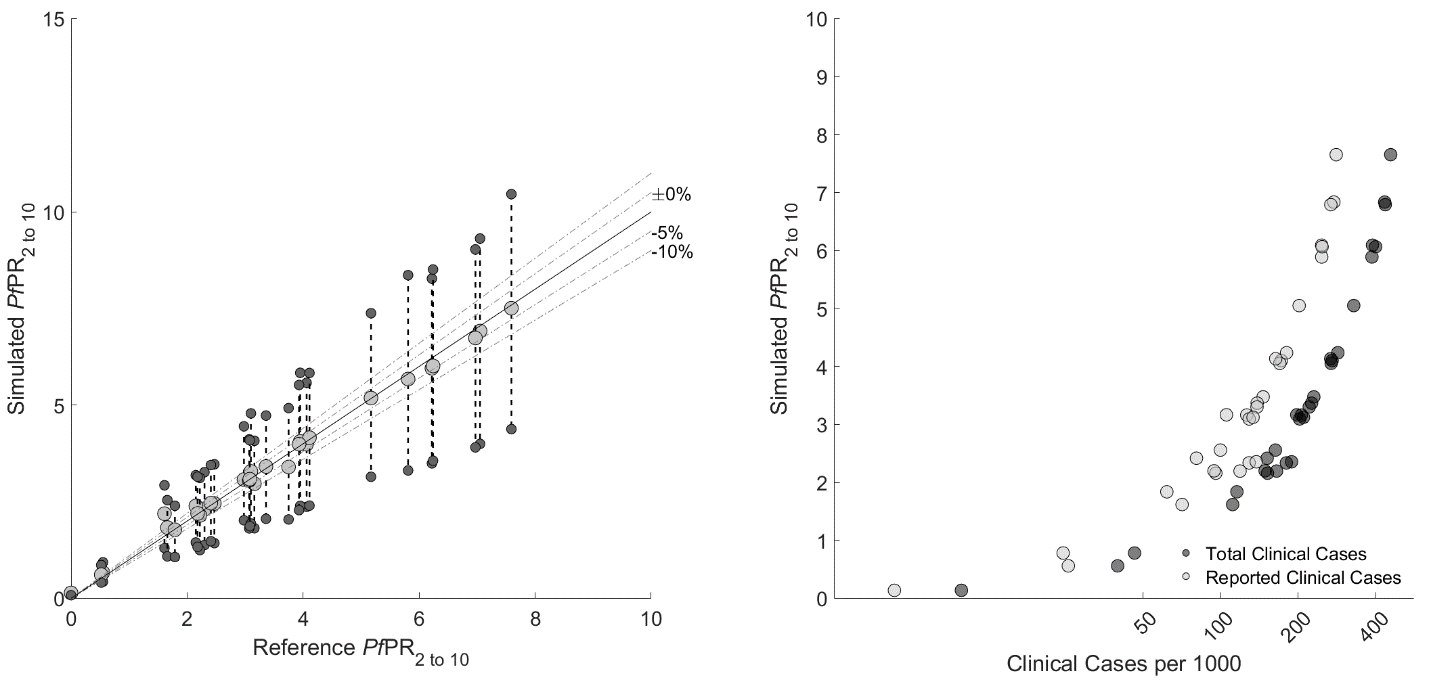

Three metrics were used for model calibration: the population weighted, district-level, annual mean *Pf*PR_2-10_ as projected by the Malaria Atlas Project (Weiss et al., 2019) versus the simulated *Pf*PR_2-10_, the district level clinical cases projected by the simulation, and the projected 561H frequency. The target deviation of simulated *Pf*PR_2-10_ versus the reference is ±10%, although for low prevalence districts, reaching this target is challenge (Figure S3). However, the overall *Pf*PR_2-10_ is acceptable, with low prevalence districts having a simulated *Pf*PR_2-10_ that skews slightly higher than the reference, whereas the higher prevalence districts skew slightly lower.

The next calibration point is a comparison of the district level projections for clinical cases versus the expected incidence for 2017 (U.S. President’s Malaria Initiative, 2019). Starting with the total clinical cases per 1000 (Figure S4), it is clear that cases are reasonably distributed in a manner that is consistent with the reference materials; although the counts of both all clinical cases, and treated cases (Figure S5) is low than that for 2017. However, the overall trend for Rwanda has been for a decline in malaria cases (U.S. President’s Malaria Initiative, 2020), so an under projection of cases by the simulation is acceptable and the projections are a reasonable. Likewise, the good agreement between the projected *Pf*PR_2-10_ and reference values supports model calibration as being within acceptable bounds for projections to be conducted.

Figure S4. Annual district level simulated clinical cases per 1000, as determined by counting all individuals within the simulation that have clinical symptoms. Map prepared by the authors using ArcGIS Pro (version 3.0) using administrative brounderies from World Bank Group (2018, 2020), geographic data from Rwanda National Institute of Statistics (2018), and prevalence data from the Malaria Atlas Project (Weiss et al., 2019).

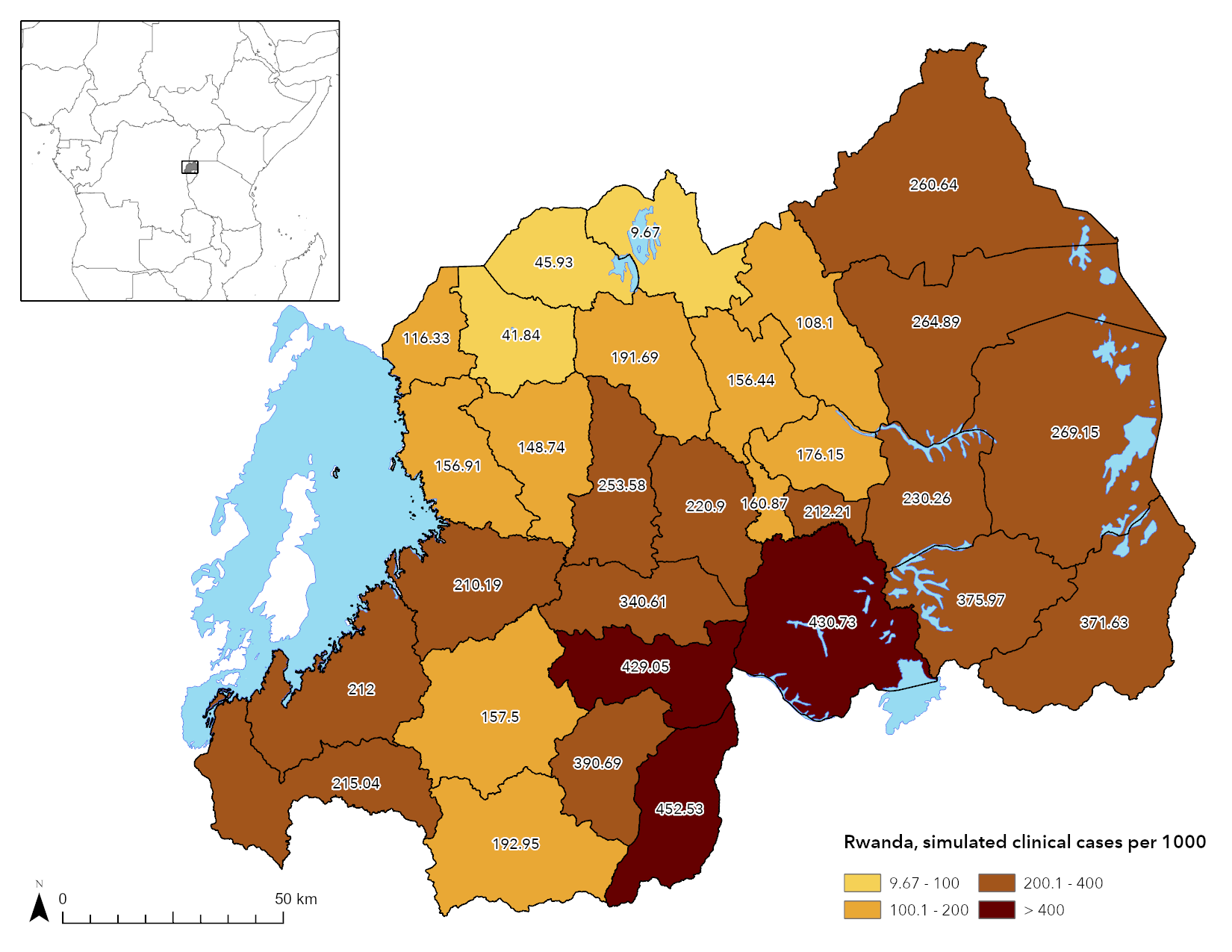

Figure S5. Annual simulated treated cases per 1000, as determined by counting all individuals that sought treatment within a given year. Map prepared by the authors using ArcGIS Pro (version 3.0) using administrative brounderies from World Bank Group (2018, 2020), geographic data from Rwanda National Institute of Statistics (2018), and prevalence data from the Malaria Atlas Project (Weiss et al., 2019).

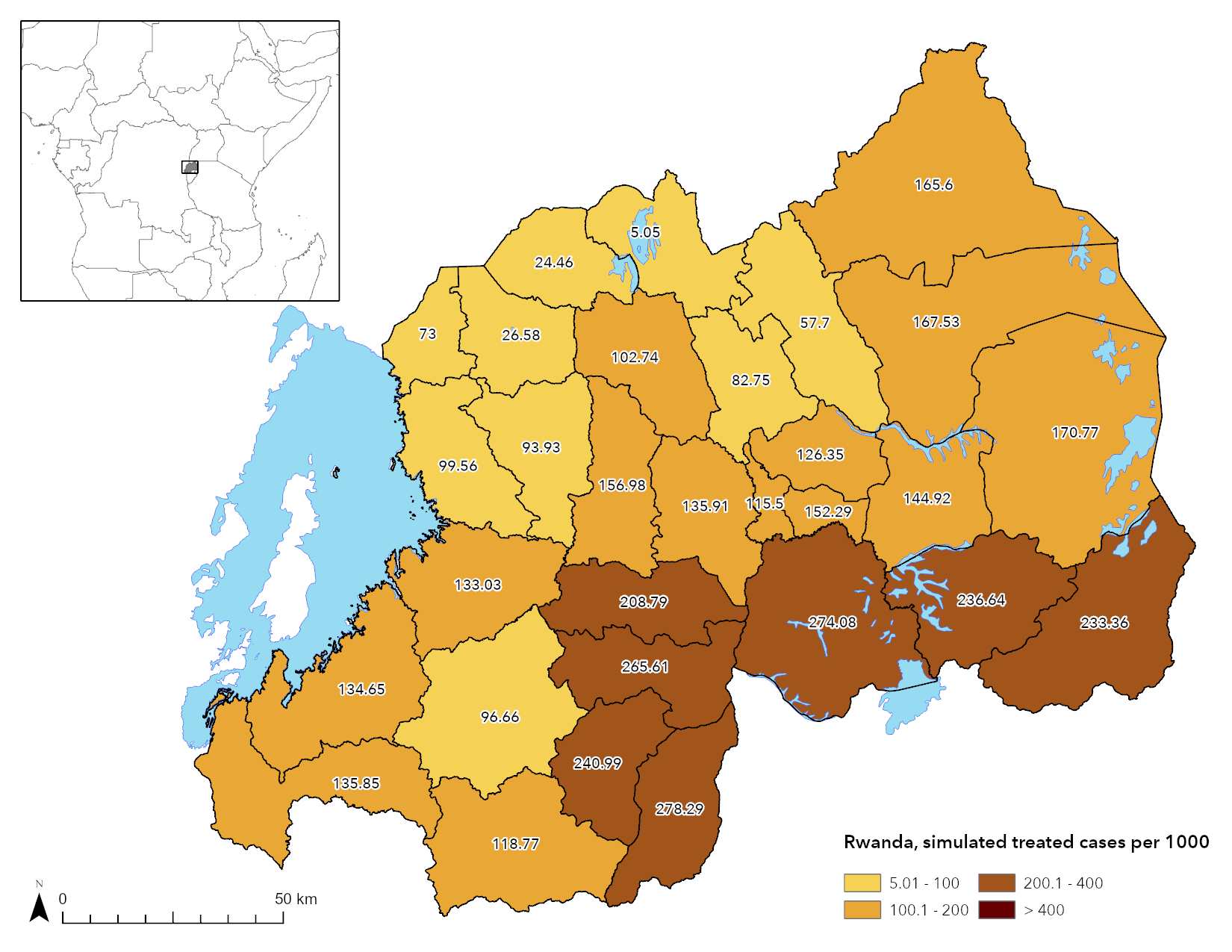

Figure S6. Spaghetti plot of replicates (*n* = 50) performed to evaluate the model calibration for the introduction of the 561H mutation. Note that at a national level (top) the projected frequency tends to be fairly smooth, although at a district level (middle, bottom) more variance is observable in the induvial replicates. Across all three plots, note that low frequencies of 561H are still possible.

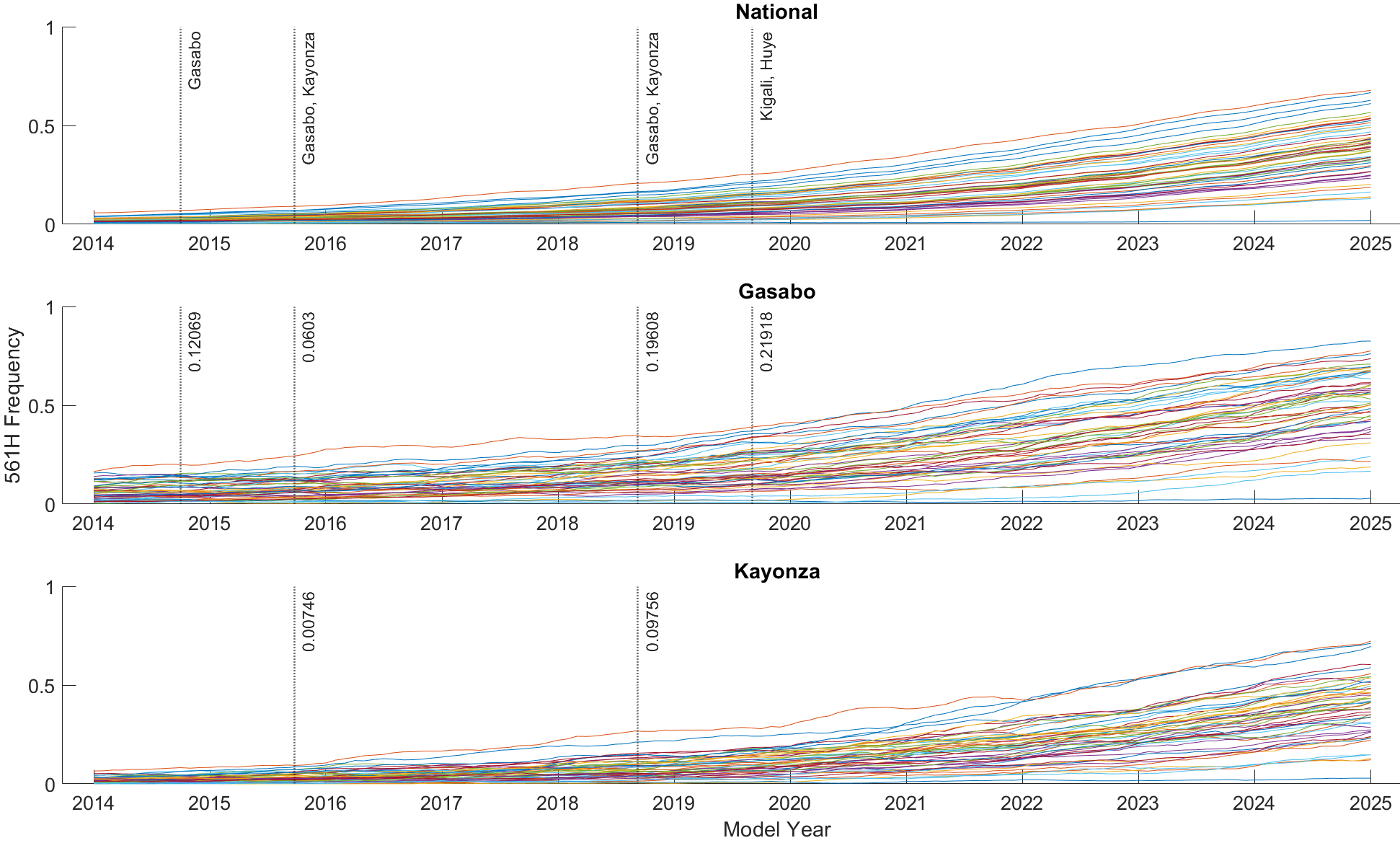

The final point for the calibration was the 561H frequency. While data is available concerning the 561H frequency in Gasabo, Kayonza, and Huye districts (Supplemental Material 2, Table S2), broad national-level frequency data is not available. As a result, the model was calibrated to utilize as single introduction in which infected individuals in Gasabo have their parasites switch from R561 to 561H. The precise fraction of individuals to have this induced mutation was determined using a parameter space search and is the only introduction of 561H in the simulation. While this introduction results in generally consistent spread of the mutation, outliners are still possible (Figure S6). The spread of the mutations with frequencies that that are similar previously found (Bergmann et al., 2021; Straimer et al., 2021; Uwimana et al., 2020, 2021), support the model as having the 561H introduction properly calibrated. However, as a note of caution, while this calibration suggests a mechanism through which 561H may have spread in Rwanda (i.e., via introduction into Gasabo) as the model was not designed, configured, or calibrated to explore this, such a conclusion should not be drawn on the basis of this work.

#### 3.2 Drug Efficacy

As part of the model calibration and validation, the drug efficacies were calculated to ensure they were within the bounds of previous studies (Nguyen et al., 2021). The complete protocol for assessing drug efficacy is elucidated in Nguyen et al. (2021); but may be summarized as follows. Within the simulation, a population of 50,000 individuals with a population distribution consistent with that of Rwanda, were inflicting *Plasmodium falciparum* infections with relevant drug resistance genotype profiles upon a population of 50,000 individuals with a. Following transition of the infection from the liver stage to the blood stage (i.e., clinical symptoms), the relevant therapy was given to the individual and the efficiency as assessed using the parasite density 28 days after the first treatment. Individuals with a parasite density less than 10 per microliter of blood were counted as cleared, failed otherwise. The complete drug efficacies are included as tables in Supplemental Materials 2.

### 4 References

Bergmann, C., van Loon, W., Habarugira, F., Tacoli, C., Jäger, J., Savelsberg, D., Nshimiyimana, F., Rwamugema, E., Mbarushimana, D., Ndoli, J., Sendegeya, A., Bayingana, C., & Mockenhaupt, F. (2021). Increase in Kelch 13 Polymorphisms in Plasmodium falciparum, Southern Rwanda. *Emerging Infectious Disease Journal*, *27*(1), 294. https://doi.org/10.3201/eid2701.203527

Malaria and Other Parasitic Diseases Division of the Rwanda Biomedical Center Ministry of Health [Rwanda] & ICF. (2018). *Rwanda Malaria Indicator Survey (RMIS) 2017* (p. 158) [MIS Final Reports]. MOPDD and ICF. https://dhsprogram.com/publications/publication-MIS30-MIS-Final-Reports.cfm

National Institute of Statistics of Rwanda. (2021). *Rwanda Vital Statistics Report—2020* (p. 86). National Institute of Statistics of Rwanda. http://www.statistics.gov.rw/publication/1705

National Institute of Statistics of Rwanda, Ministry of Health, & The DHS Program. (2020). *Rwanda Demographic and Health Survey 2019-20: Key Indicators* (p. 53) [Statistical report]. National Institute of Statistics of Rwanda. https://www.statistics.gov.rw/publication/demographic-and-health-survey-20192020-key-indicators

Nguyen, T. D., Tran, T. N.-A., Parker, D. M., White, N. J., & Boni, M. F. (2021). Antimalarial mass drug administration in large populations and the evolution of drug resistance. *BioRxiv*, 2021.03.08.434496. https://doi.org/10.1101/2021.03.08.434496

Republic of Rwanda, Ministry of Health. (n.d.). *Rwanda Health Sector Performance Report 2017-2019* (pp. 1–95). Ministry of Health. Retrieved October 13, 2020, from https://moh.gov.rw/fileadmin/Publications/Reports/FINAL%20Annual%20Report%202017-2019%2002062020.pdf

Rwanda National Institute of Statistics. (2018). *Rwanda—Water Bodies (Lakes)* [Map]. OCHA Regional Office for Southern and Eastern Africa (ROSEA). https://data.humdata.org/dataset/rwanda-water-bodies

Straimer, J., Gandhi, P., Renner, K. C., & Schmitt, E. K. (2021). High Prevalence of Plasmodium falciparum K13 Mutations in Rwanda Is Associated With Slow Parasite Clearance After Treatment With Artemether-Lumefantrine. *The Journal of Infectious Diseases*, *jiab352*. https://doi.org/10.1093/infdis/jiab352

United Nations, Department of Economic and Social Affairs, Population Division. (2019). *World Population Prospects 2019: Volume II: Demographic Profiles* (ST/ESA/SER.A/427; pp. 1–1238). United Nations. https://population.un.org/wpp/Publications/Files/WPP2019_Volume-II-Demographic-Profiles.pdf

U.S. President’s Malaria Initiative. (2019). *Rwanda Malaria Operational Plan FY 2019* (p. 58). https://www.pmi.gov/where-we-work/rwanda/

U.S. President’s Malaria Initiative. (2020). *Rwanda Malaria Operational Plan FY 2020* (p. 103). https://www.pmi.gov/where-we-work/rwanda/

Uwimana, A., Legrand, E., Stokes, B. H., Ndikumana, J.-L. M., Warsame, M., Umulisa, N., Ngamije, D., Munyaneza, T., Mazarati, J.-B., Munguti, K., Campagne, P., Criscuolo, A., Ariey, F., Murindahabi, M., Ringwald, P., Fidock, D. A., Mbituyumuremyi, A., & Menard, D. (2020). Emergence and clonal expansion of in vitro artemisinin-resistant Plasmodium falciparum kelch13 R561H mutant parasites in Rwanda. *Nature Medicine*. https://doi.org/10.1038/s41591-020-1005-2

Uwimana, A., Umulisa, N., Venkatesan, M., Svigel, S. S., Zhou, Z., Munyaneza, T., Habimana, R. M., Rucogoza, A., Moriarty, L. F., Sandford, R., Piercefield, E., Goldman, I., Ezema, B., Talundzic, E., Pacheco, M. A., Escalante, A. A., Ngamije, D., Mangala, J.-L. N., Kabera, M., … Lucchi, N. W. (2021). Association of Plasmodium falciparum kelch13 R561H genotypes with delayed parasite clearance in Rwanda: An open-label, single-arm, multicentre, therapeutic efficacy study. *The Lancet Infectious Diseases*, *21*(8), 1120–1128. https://doi.org/10.1016/S1473-3099(21)00142-0

Weiss, D. J., Lucas, T. C. D., Nguyen, M., Nandi, A. K., Bisanzio, D., Battle, K. E., Cameron, E., Twohig, K. A., Pfeffer, D. A., Rozier, J. A., Gibson, H. S., Rao, P. C., Casey, D., Bertozzi-Villa, A., Collins, E. L., Dalrymple, U., Gray, N., Harris, J. R., Howes, R. E., … Gething, P. W. (2019). Mapping the global prevalence, incidence, and mortality of Plasmodium falciparum, 2000–17: A spatial and temporal modelling study. *The Lancet*, *394*(10195), 322–331. https://doi.org/10.1016/S0140-6736(19)31097-9

World Bank Group. (2018). *Rwanda Districts* [Map]. The World Bank. https://datacatalog.worldbank.org/search/dataset/0041453

World Bank Group. (2020). *World Boundaries GeoDatabase* [GeoDatabase]. The World Bank. https://datacatalog.worldbank.org/search/dataset/0038272/World-Bank-Official-Boundaries

1. Source code available on <https://github.com/bonilab/malariaibm-spatial-Rwanda-561H> under the Source/Analysis directory.
